# Association of Prior COVID-19 Infection with Restorative Sleep Quality Using REST-Q Scores: Findings from the COPE Initiative

**DOI:** 10.1101/2025.10.16.25338136

**Authors:** Monisha Das Ireland, Mark É Czeisler, Mark E. Howard, Rebecca Robbins, Lauren A. Booker, Melinda L. Jackson, Christine F McDonald, Prerna Varma, Matthew D. Weaver, Shantha M.W. Rajaratnam, Charles A. Czeisler, Stuart F Quan

## Abstract

**Objectives:** The COVID-19 pandemic has disrupted sleep health globally. However, the relationship between prior SARS-CoV-2 infection and subjective restorative sleep, measured using validated instruments, remains underexplored. This study evaluated the association between prior SARS-CoV-2 infection and restorative sleep quality using Restorative Sleep Questionnaire (REST-Q) scores in a large, nationally representative U.S. sample.

**Methods:** Data were obtained from the September–October 2022 wave of the COVID-19 Outbreak Public Evaluation (COPE) Initiative, a cross-sectional online survey of 4,982 adults approximating the U.S. population by age, sex, race, and ethnicity. Restorative sleep was measured using the 9-item REST-Q. COVID-19 infection status was self-reported and categorized as never infected, one infection, or ≥ 2 infections. General linear models and ordinal logistic regression assessed associations between infection status and REST-Q scores, adjusting for demographic, socioeconomic, comorbidity, sleep-related, and mental health variables (Patient Health Questionnaire-4).

**Results:** Participants with prior COVID-19 infection had significantly lower REST-Q scores compared with those without infection (52.1±22.5 vs. 57.9±24.4, p<0.001). Prior COVID-19 infection was also associated with higher odds of reporting low restorative sleep (adjusted OR=1.15, 95% CI: 1.02–1.30, p=0.019). REST-Q scores decreased with increasing number of infections. In fully adjusted models including anxiety and depression, the association attenuated (p=0.078) but remained significant in sensitivity analyses accounting for infection recency.

**Conclusions:** Prior COVID-19 infection is associated with reduced restorative sleep quality as measured by REST-Q scores, independent of multiple confounders. The association persists although is partly attenuated when adjusted for anxiety and depression. These findings suggest a potential long-term impact of COVID-19 on self-reported restorative sleep and highlight the need for mechanistic and interventional research to address post-COVID sleep impairment.

**Brief Significance Statement:** This study is the first to examine the association between prior COVID-19 infection and restorative sleep using the validated Restorative Sleep Questionnaire (REST-Q) in a large, nationally representative U.S. sample. Findings demonstrate that individuals with a history of COVID-19 report significantly poorer restorative sleep, independent of multiple demographic, socioeconomic, and sleep-related factors, underscoring the need for targeted post-COVID sleep interventions.

## Introduction

The COVID-19 pandemic has profoundly impacted sleep health worldwide in a multifaceted manner. Beyond the acute respiratory and systemic manifestations of SARS-CoV-2 infection, accumulating evidence highlights a high prevalence of both acute and persistent sleep disturbances among affected individuals. In a large U.S.-based cohort, insomnia symptoms and poor sleep quality were associated with an increased risk of COVID-19 infection and hospitalization, underscoring a potential bidirectional relationship between sleep disturbance and infectious disease outcomes [1].

Longitudinal and cross-sectional studies have further illustrated the sustained impact of COVID-19 on sleep. Data from the Sleep Quest study in the United Kingdom revealed that sleep quality, timing, and duration impairments persisted well beyond the resolution of lockdown measures, suggesting a long-lasting disruption of circadian and sleep regulatory systems [2]. Similarly, a recent cross-sectional analysis reported that both acute COVID-19 and post-acute sequelae of SARS-CoV-2 infection (PASC, long COVID) were independently associated with higher rates of sleep-onset insomnia, sleep maintenance difficulties, non-restorative sleep, and impaired daytime performance [3]. Mental health conditions and primary sleep disorders, as well as psychological stressors during the pandemic, such as social isolation, financial stress, and fear of illness, can worsen sleep and thus may have resulted in less restorative sleep [4,5].

Restorative sleep, defined as the subjective experience of feeling mentally and physically refreshed upon awakening, is a critical yet underassessed domain of sleep health [6,7]. Assessing restorative sleep offers greater insight into the functional impact of sleep quality and may identify health risks not captured by duration alone [8]. Most widely used instruments, such as the Pittsburgh Sleep Quality Index (PSQI) [9], emphasize sleep disturbances and duration but do not comprehensively evaluate the restorative value of sleep. To address this gap, Robbins et al. developed the Restorative Sleep Questionnaire (REST-Q), a validated 9-item measure assessing multiple dimensions of post-sleep restoration [10]. The instrument has been shown to correlate positively with established metrics of sleep quality and duration and inversely with sleep initiation and maintenance difficulties, supporting its construct validity in a nationally representative U.S. sample.

Despite emerging literature on post-COVID sleep disruption, no prior studies have examined the relationship between COVID-19 infection and subjective restorative sleep using a validated instrument such as the REST-Q. The present study evaluates the association between prior COVID-19 infection and REST-Q scores in a general population sample. We hypothesize that, compared to individuals without such a history, individuals with a history of COVID-19 will exhibit significantly lower REST-Q scores, indicative of impaired subjective restoration.

## Methods

The study was conducted using data from Phase II of the COVID-19 Outbreak Public Evaluation (COPE) Initiative, a series of cross-sectional surveys designed to assess psychological, behavioral, and social health during the COVID-19 pandemic. These surveys were administered in 5 successive waves from March 10, 2022, to October 15, 2022. Each wave included approximately 5,000 unique participants who were recruited to reflect population estimates for age, sex, race, and ethnicity based on the 2020 U.S. Census. Questions concerning restorative sleep were asked on the 5^th^ wave (September 26-October 15, 2022) and form the basis of this analysis. Surveys were conducted online by Qualtrics, LLC (Provo, Utah, and Seattle, Washington, U.S.) using their network of participant pools with varying recruitment methodologies that include digital advertisements and promotions, word of mouth and membership referrals, social networks, television and radio advertisements, and offline mail-based approaches. Informed consent was obtained electronically. The study was approved by the Monash University Human Research Ethics Committee (Study #24036).

### Survey Items

Participants self-reported their demographic, anthropometric, and socio-economic information, including age, race, ethnicity, sex, height and weight, education level, employment status, and household income. They also provided details on the number of COVID-19 infections they had experienced and their COVID-19 vaccination status.

Ascertainment of past COVID-19 infection was obtained using responses from the following questions related to COVID-19 testing or the presence of loss of taste or smell:

1. Have you ever tested positive?
2. Despite never testing positive, are you confident that you have had COVID-19?
3. Despite never testing positive, have you ever received a clinical diagnosis of COVID-19?
4. Have you experienced a problem with a decreased sense of taste or smell at any point since January 2020?

Restorative sleep quality was assessed using the validated REST-Q, which consists of 9 self-reported items worded to reflect both positive and negative experiences immediately upon awakening [10]. Participants were asked, “For each of the following items, please tell me to what degree you feel each of the below when you woke up today, compared to before you went to sleep”. Last night’s sleep left me feeling “tired”, “sleepy”, “in a good mood”, “rested”, “refreshed”, “ready to start the day”, “energetic”, “mentally alert”, “grouchy”. Each item is rated on a 5-point Likert scale, where 0 = "Not at all," 1 = "Rarely," 2 = "Sometimes," 3 = "Often," and 4 = "Always." The items “tired”, “sleepy,” and “grouchy” were reverse-coded. Responses from the 9 REST-Q questions were then converted into a 100-point scale. A score of 50 and below is categorized as a “low” REST-Q score, a score ranging from 50 to 74.99 is categorized as a “somewhat” REST-Q score, and a score of 75 and above is categorized as a “high” REST-Q score.

Questions regarding sleep duration and disturbances included: “During the past month, how many hours of actual sleep did you get at night?” and “How many nights in a week do you have problems with your sleep?” Data concerning insomnia symptoms, sleep quality, and obstructive sleep apnea (OSA) were obtained from responses to the PSQI. The Patient Health Questionnaire-4 (PHQ-4) was used to screen for anxiety and depression [11].

Participants were considered to have OSA if they endorsed currently having the condition, whether treated or not, or if they had two or more symptoms of OSA (i.e., snoring, breathing pauses, or sleepiness) on the PSQI. Insomnia was present if endorsed by participants, irrespective of treatment status. Trouble with poor sleep was present if its extent was “Much” or “Very Much”. Poor sleep quality was defined as being endorsed as “Fairly Bad” or “Very Bad”.

Participants reported on co-morbid conditions by answering the question, “Have you been newly diagnosed with any of the following medical conditions since January 2020?” These conditions included high blood pressure, cardiovascular disease (eg, heart attack, stroke, angina), gastrointestinal disorder (e.g., acid reflux, ulcers, indigestion), cancer, chronic kidney disease, liver disease, sickle cell disease, chronic obstructive pulmonary disease and asthma.

### Statistical Analyses

Summary data for continuous variables are reported as their respective means and standard deviations (SD) or standard errors (SE), and for categorical and ordinal variables as their percentages [10]. We defined predictive variables consistent with prior analyses of the COPE cohort [12]. A positive history of COVID-19 infection was defined as an affirmative response to having tested positive for COVID-19 or a new loss of taste or smell. We collapsed the number of COVID-19 infections into an ordinal variable with three levels (never infected, one infection, two or more infections). The number of COVID-19 vaccinations was dichotomized as boosted (>2 vaccinations) or not boosted (≤2 vaccinations). Comorbid medical conditions were defined as currently having the condition, whether treated or untreated. The effect of comorbid medical conditions was evaluated by summing the number of conditions reported by the participant (minimum value 0, maximum value 9). Sleep duration was collapsed into an ordinal variable with three levels (<5 hours, 5-8 hours, >8 hours). Socioeconomic covariates were dichotomized as follows: employment (full-time, part-time, student vs. not employed), education (high school or less vs. some college or higher), and income in U.S. Dollars to approximate 200% of the 2022 U.S. Poverty Level for a family of 4 (<$50,000 vs ≥$50,000).

Comparisons of REST-Q scores among levels of categorical or ordinal variables were performed using Student’s unpaired t-test or analysis of variance. Bivariate comparisons of REST-Q scores between continuous variables were conducted by calculating Pearson correlation coefficients. Additionally, REST-Q scores were categorized as “Low”, “Somewhat,” and “High” according to the original validation of the instrument with bivariate comparisons made using χ^2^ [10].

Missing data were infrequent (3.5% of the observations). However, inclusion of participants with complete data would have result in the exclusion of 51.2% of the cases. Therefore, multiple imputation by chained equations was employed to generate replacement values. Comparison of the imputed dataset with the original dataset revealed no outliers in the imputed dataset, and the means of the same variables between the two datasets were comparable.

General linear modeling using the imputed dataset was employed to determine whether there was a difference in REST-Q scores as a function of COVID-19 infection after adjusting for relevant covariates. To reduce the number of relevant predictors, minimize overfitting of models, diminish potential collinearity, and minimize prediction error, we performed an elastic net regression on the original dataset using 10-fold cross-validation to select the covariates with non-zero coefficients that might contribute to an overall model of the REST-Q score. We then developed increasingly complex models by sequentially including demographic and socioeconomic factors, comorbidity, sleep factors, and mental health conditions. Similarly, progressively complex multiple ordinal logistic regression models were used to determine whether categories of REST-Q score were associated with COVID-19 infection. Sensitivity analyses were conducted by using more stringent (positive COVID-19 test only) and less stringent (positive COVID-19 test, loss of taste or smell, or clinical diagnosis) definitions of COVID-19 infection. In addition, to assess the impact of the interval between a participant’s last COVID-19 infection and the timing of the COPE survey, analyses were performed by incorporating this interval as a covariate. To determine the impact of recent COVID-19 infection, we repeated analyses by excluding participants for whom the interval was ≤ 30 days.

All analyses were conducted using SPSS version 28 (IBM, Armonk, NY). A p<0.05 was considered statistically significant.

## Results

Characteristics of the 4982 participants are shown in Table 1. Mean age of the cohort was 47.3±17.5 years. There were slightly more women in the sample (51.4%) and most participants identified as Non-Hispanic White. Prior COVID-19 infection was reported by 2246 (45.1%) participants. The mean interval between last COVID-19 infection and survey date was 377.1±280.5 days. The prevalences of Low, Somewhat and High category REST-Q scores were 39.0%, 36.4% and 24.5%, respectively.

Table 2 shows univariate associations between REST-Q scores and participant characteristics. The overall mean REST-Q score was 55.3±23.8. Participants with a history of COVID-19 infection had lower REST-Q scores than those without infection (52.1±22.5 vs. 57.9±24.4, p<0.001). REST-Q scores declined with increasing number of infections, and participants who received a COVID-19 booster had higher scores than those who did not (57.4±23.0 vs. 52.3± 23.8, p <0.001). Higher REST-Q scores were also observed in those who were older, were women, had higher income, lived with a partner, had higher educational attainment, and were not employed. No differences were noted related to BMI or number of comorbidities. Except for those who were Other Non-Hispanic, little variation was seen among combined race and ethnicity groups. Participants who endorsed having problems with their sleep, insomnia, and symptoms consistent with obstructive sleep apnea had lower REST-Q scores. As self-reported sleep duration declined, REST-Q scores decreased as well. Participants with anxiety or depression symptoms (PHQ-4) had significantly lower REST-Q scores compared to those without symptoms (Anxiety: 41.2±20.5 vs. 59.7±22.9, p<0.001; Depression: 40.9±20.4 vs. 59.9±22.9, p <0.001).

Table 3 presents the association between REST-Q scores and self-reported COVID-19 infection status after adjusting for relevant covariates. The baseline model, unadjusted for covariates, indicated that the COVID-19 negative group had significantly higher REST-Q scores (57.9±0.5) than the COVID-19 positive group (52.1±0.5; p < .001). This relationship remained consistent after adjusting for potential confounders with increasingly complex models. In the fully adjusted model, which included anxiety and depression along with all previous covariates, the difference in REST-Q scores was attenuated, but still approached statistical significance. (COVID-19 positive group [47.5±0.7] and COVID-19 negative group [48.5±=0.7; p=0.078].

Also shown in Table 3 is the effect of the interval between the last COVID-19 infection and the date of the survey on the association of REST-Q scores and COVID-19 infection in the model. Addition of this interval as a covariate in the last model still found that COVID-19 infection was associated with lower REST-Q scores (46.4±1.2 vs. 48.7±1.4, p=0.021). This relationship was maintained even after exclusion of participants (n=94) in whom this interval was ≤ 30 days (46.5±1.3 vs. 48.8±1.4, p=0.019).

Table 4 presents the results of ordinal logistic regression models assessing the association between COVID-19 infection status and categories of REST-Q scores. In the baseline unadjusted model, individuals with a history of COVID-19 infection had significantly higher odds of reporting worse restorative sleep compared to those without prior infection (OR = 1.561; 95% CI: 1.407–1.731; p < .001). This association remained statistically significant across sequential models adjusting for demographics (OR = 1.264; 95% CI: 1.135–1.411; p < .001), socioeconomic factors (OR = 1.329; 95% CI: 1.191–1.484; p < .001), comorbidities (aOR = 1.286; 95% CI: 1.149–1.439; p < .001), sleep characteristics (OR = 1.202; 95% CI: 1.068–1.354; p = .002), and anxiety and depression (PHQ-4; fully adjusted model: OR = 1.153; 95% CI: 1.024–1.301; p = .019).

Table 5 presents the results of sensitivity analyses assessing the robustness of the association between COVID-19 infection status and REST-Q scores, stratified by two alternative definitions of COVID-19 positivity. When COVID-19 positivity was defined solely as a self-reported positive COVID-19 test, the estimated mean REST-Q score was lower (47.1±0.7) in the COVID-19 positive group compared to (48.5±0.7) the COVID-19 negative group (p < .019; Table 5A). In a fully adjusted ordinal regression model with three categories of REST-Q scores, there also was a significant association of lower REST-Q scores with COVID-19 defined only as a positive test (aOR: 1.166, 95% CI: 1.034-1.314, p = 0.012) (Table 5B). In the second analysis, COVID-19 positivity was defined more liberally as a self-reported positive COVID-19 test, loss of taste or smell, or a clinical diagnosis of COVID-19. Under this definition, the mean REST-Q score for the positive group (47.7±0.7) was less than for the negative group (48.3±0.7, p < .012) (Table 5A). In a fully adjusted ordinal regression model, the more liberal definition that included clinical diagnosis of COVID-19 infection or a loss of taste or smell still showed a trend towards association, although it was not statistically significant (aOR: 1.104, 95% CI: 0.979-1.245, p = 0.105) (Table 5B).

## Discussion

In this large, cross-sectional analysis of nearly 5,000 U.S. adults, we found that individuals with a history of COVID-19 infection reported significantly poorer restorative sleep quality, as assessed by the validated REST-Q instrument. A dose-response relationship was observed, with REST-Q scores declining progressively as a function of increasing number of COVID-19 infections. Furthermore, participants who received a COVID-19 booster exhibited higher REST-Q scores than those who did not, suggesting a possible protective effect of vaccination. While the association between COVID-19 and REST-Q scores attenuated after controlling for mental health symptoms (PHQ-4), the relationship between COVID-19 and poor restorative sleep nonetheless persisted.

Our finding that COVID-19 infection was associated with less restorative sleep is consistent with a growing body of research demonstrating post-COVID-19 sleep disturbance as a prevalent and persistent symptom. Abuhammad et al. reported that 40.6% of recovered COVID-19 patients experienced poor sleep quality, while 63.6% reported sleep pattern disturbances during the post-COVID era. Their findings also highlighted socioeconomic and demographic predictors of poor sleep, including younger age and lower income, which mirror trends observed in our cohort [13]. Tedjasukmana et al. highlighted sleep disturbances in up to 78% of patients post-infection, including insomnia, sleep-disordered breathing, and disruptions to circadian rhythm, with detrimental effects on quality of life [14]. Similarly, Munteanu et al. found that over half of their Romanian cohort reported poor sleep quality (PSQI ≥5) as long as 6 months after infection, regardless of initial disease severity [15]. However, these studies primarily focused on sleep quantity and quality rather than subjective sleep restoration.

Restorative sleep, defined as the subjective feeling of being mentally and physically refreshed upon waking, is a vital yet underexplored aspect of sleep health. Most common tools, like the PSQI, focus on sleep disturbance and duration but do not provide a comprehensive measure of sleep’s restorative quality. Despite increasing research on post-COVID sleep issues [13–16], no previous studies have systematically investigated the link between COVID-19 infection and subjective restorative sleep using a validated tool such as the REST-Q. Our study is the first to examine this relationship in a large, diverse population sample. As we hypothesized, we found that individuals with a history of COVID-19 scored significantly lower on the REST-Q scores, indicating reduced subjective restoration compared to those without prior infection.

In univariate analyses, we found that anxiety, depression, insomnia, OSA, and reports of sleep problems and poor sleep quality were associated with diminished restorative sleep. In particular, anxiety and depression have been shown to have strong, bidirectional relationships with sleep disturbance [17,18] and could have accounted for the preponderance of the observed effects in our study. While prior studies have consistently demonstrated this bidirectional association between non-restorative sleep and mental health outcomes, no studies to date have specifically described a relationship between restorative sleep and mental health, highlighting a gap in the literature. Furthermore, the association between COVID-19 infection and diminished restorative sleep persisted after controlling for several socio-economic, medical, and mental health factors, including anxiety and depression. This indicates that our findings cannot be explained solely on external features and suggests that they are related to an independent effect of the infection on the neurobiology of sleep.

Several potential mechanisms may explain our findings. COVID-19 infection can result in SARS-CoV-2 neurotropism affecting sleep-regulating brain regions. In a recent brain magnetic resonance imaging study of persons after COVID-19 infection, worse sleep quality was associated with a smaller putamen volume [19]. In addition, COVID-19 infection can lead to persistent inflammatory responses (e.g., elevated IL-6, TNF-α), mitochondrial dysfunction, and dysregulation of the autonomic nervous system [20–22]. These consequences of infection have been associated with sleep disturbances.

This study’s primary strength lies in its large, demographically representative sample of U.S. adults and the breadth of covariate data. The inclusion of sociodemographic, medical, mental health, and sleep-related variables enabled detailed multivariable adjustment and stratified analyses, which few prior studies on post-COVID sleep outcomes have achieved. Sensitivity analyses using alternative definitions of COVID-19 exposure further strengthened the robustness of our findings. Furthermore, exclusion of participants who recently experienced COVID-19 infection and who may have been experiencing sleep disturbance from their acute illness did not mitigate our results.

However, several limitations must be acknowledged. The cross-sectional design precludes causal inference, limiting our ability to determine whether COVID-19 infection directly causes impaired restorative sleep or whether reverse causality occurred. Another limitation lies in the reliance on self-reported data for both COVID-19 history and sleep measures. While the REST-Q is a validated tool, self-reported sleep metrics are inherently susceptible to recall bias and may not fully capture objective sleep architecture or efficiency.

### Conclusion

This study examined the relationship between COVID-19 infection and restorative sleep quality using data from the COPE Initiative and a validated instrument, the REST-Q questionnaire. Through multiple analytic approaches, COVID-19 infection was consistently linked to reduced restorative sleep. The consistency of results across various statistical models and varying definitions of COVID-19 exposure provides strong evidence of an association between prior COVID-19 infection and reduced restorative sleep. These results highlight the potential long-term effects of COVID-19 on subjective sleep recovery and underscore the need for further research to understand the underlying mechanisms and identify potential interventions. Given the attenuation of the association between COVID-19 and REST-Q scores after adjusting for anxiety and depression, future studies should explore the mediating role of mental health symptoms and their effect on sleep quality in post-COVID-19 patients.

## Statements and Declarations

### Ethics Approval

All procedures were in accordance with the ethical standards of Monash University Human Research Ethics Committee (Study #24036) and with the 1964 Helsinki declaration and its later amendments or comparable ethical standards. Informed consent was obtained electronically from all individual participants included in the study.

### Author Approval and Consent to Publish

All authors have seen and approved the manuscript; all authors have given their consent to publish.

## Acknowledgments of Author Contributions

Concept and Design: MDI, SFQ

Data collection: MDW, MÉC, MEH

Data analysis and interpretation: SFQ

Drafting of the manuscript: MDI, SFQ

Critical feedback and revision of manuscript: MDI, SFQ, MDW, MÉC, LAB, MEH, MLJ, CFM, RR, PV, SMWR, CAC

## Data Availability

The data that support the findings of this study are available from the corresponding author upon reasonable request subject to any institutional review board constraints.

