## Supplementary material for "Association of Prior COVID-19 Infection with Restorative Sleep Quality Using REST-Q Scores: Findings from the COPE Initiative": Table 1, Table 2, Table 3, Table 4, Table 5

| **Table 1: Participant Characteristics** |  |  |
| --- | --- | --- |
|  | *N* | *Mean (SD)/%* |
| **Overall** | 4982 |  |
| **Age** | 4982 | 47.3±17.5 |
| **Sex** |  |  |
| Men | 2405 | 48.6 |
| Women | 2547 | 51.4 |
| **Race** |  |  |
| White | 2839 | 57.0 |
| Black | 558 | 11.2 |
| Asian | 318 | 6.4 |
| Other Non-Hispanic | 312 | 6.3 |
| Hispanic | 954 | 19.2 |
| **Presence of Live In Partner** |  |  |
| No | 2071 | 41.6 |
| Yes | 2911 | 58.4 |
| **Body Mass Index (BMI, kg/m2)** | 4982 | 27.8±6.1 |
| **COVID Infection** |  |  |
| Yes | 2246 | 45.1 |
| No | 2736 | 54.9 |
| **Number of COVID-19 Infections** |  |  |
| Never Infected | 1791 | 44.8 |
| 1 Infection | 1630 | 40.8 |
| 2 or more infections | 576 | 14.4 |
| **COVID-19 Vaccine Booster Status** |  |  |
| Not Boosted | 2838 | 67.5 |
| Boosted | 1365 | 27.4 |
| **Interval Between Last COVID-19 and Survey** | 2031 | 377.1+280.5 |
| **Education** |  |  |
| High School or Less | 1210 | 24.3 |
| At least Some College | 3772 | 75.7 |
| **Employment Status** |  |  |
| Employed (Full Time/Part time) | 2462 | 58.3 |
| Not Employed or Retired | 1763 | 41.7 |
| **Income** |  |  |
| <$50,000 | 2126 | 44.3 |
| >$50,000 | 2671 | 55.7 |
| **RESTQ Scores** |  |  |
| Low | 1945 | 39.0 |
| Somewhat | 1815 | 36.4 |
| High | 1222 | 24.5 |
| Somewhat + High | 3037 | 60.9 |
| **Sleep Problems** |  |  |
| No | 4029 | 80.9 |
| Yes | 953 | 19.1 |
| **PSQI** |  |  |
| Poor Quality Sleep | 1279 | 25.7 |
| Good Quality Sleep | 3703 | 74.3 |
| **Number of Hours of Sleep** |  |  |
| < 5 hours | 1953 | 41.0 |
| 5-8 Hours | 2340 | 49.2 |
| > 8 hours | 465 | 9.8 |
| **Insomnia** |  |  |
| No | 4089 | 82.1 |
| Yes | 893 | 17.9 |
| **Obstructive Sleep Apnea Treatment** |  |  |
| No | 4036 | 81.0 |
| Yes | 946 | 19.0 |
| **Number of Comorbidities** | 4982 | 1.5±2.2 |
| **PHQ-4 Anxiety and Depression** |  |  |
| Anxiety: No | 3784 | 76.0 |
| Anxiety: Yes | 1198 | 24.0 |
| Depression: No | 3766 | 75.6 |
| Depression: Yes | 1216 | 24.4 |

| **Table 2: Univariate Associations between REST Q Scores, COVID 19 infection** | | | | |
| --- | --- | --- | --- | --- |
| **and Demographic, Anthropometric, Socioeconomic, Sleep Characteristics** | | | | |
| ***REST Q SCORES*** | | |  |  |
|  | **N** | **Mean** | **SD** | **p-valve** |
| **Overall** | 4982 | 55.3 | 23.8 |  |
| **COVID Infection** |  |  |  |  |
| Yes | 2735 | 57.9 | 24.4 | <.001 |
| No | 2246 | 52.1 | 22.5 |  |
| **RESTQ Scores** |  |  |  |  |
| Low | 1945 | 31.3 | 12.6 | <.001 |
| Somewhat | 1815 | 60.2 | 7.1 |  |
| High | 1222 | 86.1 | 8.4 |  |
| Somewhat+High | 3037 | 70.6 | 14.8 |  |
| **Age** |  |  |  |  |
| <Median (47 years) | 2506 | 49.9 | 22.2 | <.001 |
| ≥Median (47 years) | 2476 | 60.5 | 24.1 |  |
| **Sex** |  |  |  |  |
| Men | 2405 | 48.3 | 21.95 | <.001 |
| Women | 2547 | 51.1 | 24.54 |  |
| **Race** |  |  |  |  |
| White | 2839 | 55.6 | 24.3 | <.015 |
| Black | 558 | 56.5 | 23.9 |  |
| Asian | 318 | 56.2 | 21.7 |  |
| Other Non-Hispanic | 312 | 50.8 | 26.6 |  |
| Hispanic | 954 | 55.5 | 22.9 |  |
| **Body Mass Index (BMI, kg/m2)** |  |  |  |  |
| Not Overweight (<25 kg/m2) | 1674 | 55.6 | 24.3 | 0.772 |
| Overweight (≥25 kg/m2) | 2952 | 55.4 | 23.8 |  |
| **Income** |  |  |  |  |
| <$50,000 | 2126 | 51.5 | 24.8 | <.001 |
| >$50,000 | 2671 | 58.8 | 22.6 |  |
| **Presence of Live In Partner** |  |  |  |  |
| No | 2071 | 52.9 | 24.5 | <.001 |
| Yes | 2911 | 57.2 | 23.3 |  |
| **Education** |  |  |  |  |
| High School or Less | 1210 | 50.9 | 24.8 | <.001 |
| At least Some College | 3772 | 56.8 | 23.4 |  |
| **Employment Status** |  |  |  |  |
| Employed (Full Time/Part time) | 2462 | 54.7 | 21.9 | <.001 |
| Not Employed or Retired | 1763 | 59.9 | 24.8 |  |
| **Number of COVID-19 Infections** |  |  |  |  |
| Never Infected | 1791 | 57.6 | 24.7 | <.001 |
| 1 Infection | 1630 | 53.6 | 23.5 |  |
| 2 or more infections | 576 | 49.6 | 21.9 |  |
| **COVID-19 Vaccine Booster Status** |  |  |  |  |
| Not Boosted | 2838 | 52.3 | 23.8 | <.001 |
| Boosted | 1365 | 57.4 | 23.0 |  |
| **Sleep Problems** |  |  |  |  |
| No | 4029 | 59.7 | 22.2 | <.001 |
| Yes | 953 | 36.5 | 21.9 |  |
| **PSQI** |  |  |  |  |
| Poor Quality Sleep | 1279 | 62.7 | 20.7 | <.001 |
| Good Quality Sleep | 3703 | 34.6 | 19.4 |  |
| **Number of Hours of Sleep** |  |  |  |  |
| < 5 hours | 1953 | 47.5 | 22.8 | <.001 |
| 5-8 Hours | 2340 | 62.4 | 22.6 |  |
| > 8 hours | 465 | 56.8 | 23.7 |  |
| **Insomnia** |  |  |  |  |
| No | 4089 | 57.8 | 23.5 | <.001 |
| Yes | 893 | 43.9 | 22.2 |  |
| **Obstructive Sleep Apnea Treatment** |  |  |  |  |
| No | 4036 | 56.5 | 24.4 | <.001 |
| Yes | 946 | 50.1 | 20.7 |  |
| **Comorbidities** |  |  |  |  |
| No Comorbidities | 2373 | 55.6 | 24.2 | 0.267 |
| > 1 Comorbidities | 2609 | 54.9 | 23.4 |  |
| **PHQ-4 Anxiety and Depression** |  |  |  |  |
| No Distress | 2941 | 64.2 | 22.2 | <.001 |
| Mild Distress | 900 | 44.4 | 18.5 |  |
| Moderate Distress | 699 | 44.7 | 17.6 |  |
| Severe Distress | 442 | 34.8 | 23.4 |  |
| Anxiety: No | 3784 | 59.7 | 22.9 | <.001 |
| Anxiety: Yes | 1198 | 41.2 | 20.5 |  |
| Depression: No | 3766 | 59.9 | 22.9 | <.001 |
| Depression: Yes | 1216 | 40.9 | 20.4 |  |

| **Table 3: Comparison of RESTQ Scores Between COVID-19 Negative and COVID-19 Positive Status** | | | | | |
| --- | --- | --- | --- | --- | --- |
|  | **COVID-19 Negative** | | **COVID-19 Positive** | |  |
| *Model* | *Estimated Mean* | *SE* | *Estimated Mean* | *SE* | *p-value* |
| Baseline | 57.9 | 0.5 | 52.1 | 0.5 | <0.001 |
| +Demographics* | 59.4 | 0.7 | 56.5 | 0.7 | <0.001 |
| +Socioeconomic† | 59.5 | 0.7 | 56.0 | 0.7 | <0.001 |
| +Comorbidities‡ | 59.7 | 0.7 | 56.8 | 0.8 | <0.001 |
| +Sleep Factors§ | 50.7 | 0.7 | 49.2 | 0.7 | <0.05 |
| +Anxiety and Depression¶ | 48.5 | 0.7 | 47.5 | 0.7 | 0.078 |
| Effect of Interval Between Last Infection and Date of Survey | | |  |  |  |
| +Days between last infection and survey | 48.7 | 1.4 | 46.4 | 1.2 | 0.021 |
| Exclusion of participants with <30 days between last infection and survey | 48.8 | 1.4 | 46.5 | 1.3 | 0.019 |
| Models adjusted as follows: |  |  |  |  |  |
| *Age, Sex, Race |  |  |  |  |  |
| †Education, Income |  |  |  |  |  |
| ‡Number of comorbidities, Vaccination Status, BMI | |  |  |  |  |
| §Sleep Duration, Sleep Trouble, Sleep Quality | |  |  |  |  |
| ¶Anxiety and Depression |  |  |  |  |  |

| **Table 4: Association of Categories of Restorative Sleep with COVID-19 Infection** | | | | |
| --- | --- | --- | --- | --- |
|  |  | 95% CI | |  |
| *Model* | *OR/aOR* | *Lower* | *Upper* | *p-value* |
| Baseline | 1.560 | 1.406 | 1.731 | <0.001 |
| +Demographics* | 1.264 | 1.135 | 1.411 | <0.001 |
| +Socioeconomic† | 1.329 | 1.191 | 1.484 | <0.001 |
| +Comorbidities‡ | 1.286 | 1.149 | 1.439 | <0.001 |
| +Sleep Factors§ | 1.202 | 1.068 | 1.354 | 0.002 |
| +Anxiety and Depression¶ | 1.153 | 1.024 | 1.301 | 0.019 |
| Ordinal Regression Models reflect odds of a higher Rest Q category and are adjusted as follows: | | | | |
| *Age, Sex, Race |  |  |  |  |
| †Education, Income |  |  |  |  |
| ‡Number of comorbidities, Vaccination Status, BMI | | |  |  |
| §Sleep Duration, Sleep Trouble, Sleep Quality | | |  |  |
| ¶Anxiety and Depression |  |  |  |  |

| **Table 5: Sensitivity Analyses** |  |  |  |  |  |
| --- | --- | --- | --- | --- | --- |
| A: RESTQ Score Stratified by COVID-19 Status | |  |  |  |  |
|  | COVID-19 Negative | | COVID-19 Positive | |  |
| *Definition of COVID-19 Status* | *Estimated Mean* | *SE* | *Estimated Mean* | *SE* | *p-value* |
| Positive COVID-19 Test | 48.5 | 0.6 | 47.1 | 0.7 | 0.019 |
| Positive COVID-19 Test, Loss of Taste or Smell, Clinical Diagnosis | 48.3 | 0.7 | 47.7 | 0.7 | 0.330 |
| B: Ordinal Regression of Restorative Sleep Categories Stratified by COVID-19 Status | | | | |  |
|  |  | 95% CI | |  |  |
| *Definition of COVID-19 Status* | *OR/aOR* | *Lower* | *Upper* | *p* |  |
| Positive COVID-19 Test | 1.166 | 1.034 | 1.314 | 0.012 |  |
| Positive COVID-19 Test, Loss of Taste or Smell, Clinical Diagnosis | 1.104 | 0.979 | 1.245 | 0.105 |  |
